# A Risk Prediction and Clinical Guidance System for Evaluating Patients with Recurrent Infections

**DOI:** 10.1101/2020.06.12.20129692

**Authors:** Nicholas L. Rider, Gina Cahill, Tina Motazedi, Lei Wei, Ashok Kurian, Lenora M. Noroski, Filiz O. Seeborg, Ivan K. Chinn, Kirk Roberts

## Abstract

**Background:** Primary immunodeficiency diseases represent an expanding set of heterogeneous conditions which are difficult to recognize clinically. Diagnostic rates outside of the newborn period have not changed appreciably. This concern underscores a need for novel methods of disease detection.

**Objective:** We built an artificial intelligence model to provide real-time risk assessment about primary immunodeficiency and to facilitate prescriptive analytics for initiating the most appropriate diagnostic work up. Our goal is to improve diagnostic rates for primary immunodeficiency and shorten time to diagnosis.

**Methods:** We extracted data from the Texas Children’s Hospital electronic health record on a large population of primary immunodeficiency patients (n = 1762) and age-matched set of controls (n = 1698). From the cohorts, clinically relevant prior probabilities were calculated enabling construction of a Bayesian network probabilistic model. Our model was constructed with clinical-immunology domain expertise, trained on a balanced cohort of 100 cases-controls and validated on an unseen balanced cohort of 150 cases-controls. Performance was measured by area under the receiver operator characteristic curve (AUROC).

**Results:** Our Bayesian network was accurate in classifying immunodeficiency patients from controls (AUROC = 0.945; p<0.0001) at a risk threshold of ≥6%. Additionally, the model was 89% accurate for categorizing validation cohort members into appropriate International Union of Immunological Societies diagnostic categories.

**Conclusion:** Artificial intelligence methods can classify risk for primary immunodeficiency and guide management. Our Bayesian network enables highly accurate, objective decision making about risk and guides the user towards the appropriate diagnostic evaluation for patients with recurrent infections.

## Introduction

Infectious diseases contribute substantially to the costs of healthcare. For invasive Streptococcus pneumoniae alone, one estimate suggests that $3.5 billion US Dollars (USD) are spent in direct costs for complications related to this single organism.[1] Among individuals who suffer infectious disease, a subsection will have ongoing risk of morbidity and mortality owing to an underlying genetic susceptibility. Such patients with primary immune defects (PI) often present in subtle fashion and may be mistaken for individuals with routine infection leading to diagnostic delay and death.[2]

Once thought to be rare, there are now 416 distinct PI diseases with 430 molecular etiologies known.[3] However, due to improved understanding about its biology and expanded testing capabilities, PI is now estimated to affect between 1:1000 to 1:5000 individuals and our prior work suggest that upwards of 1% of the general population may have features of PI risk.[4-6] Yet, early recognition of PI remains a challenge with time to diagnosis remaining largely fixed over the past 4 decades and is estimated at 7.5 – 9 years from symptom onset.[2] Projected annual costs for PI patients prior to diagnosis is nearly $140,000 USD which may be reduced by approximately $78,000 USD following diagnosis.[7, 8] In order to spare costs and undue morbidity, efforts are underway to facilitate early PI diagnosis and treat affected patients in a precise manner.[4, 9]

To aid in disease diagnosis, artificial intelligence methods can assist in population-wide and individual-level risk assessment.[10, 11] In particular, models which meld reasoning and uncertainty can facilitate diagnosis amongst at-risk individuals.[12] Bayesian networks (BNs) are one such model which combine relevant features, joint probability distributions and an intuitive structure to answer questions in biomedicine.[13] Specifically, BNs have proven useful for clinical decision support for detecting lung cancers, prediction of heart failure, computing survival for individuals with colon cancer, aiding in liver disease evaluation and enabling pathology specimen evaluation among other use cases.[6, 14-19] It is for these reasons that we chose to build a BN for PI risk prediction and clinical guidance.

More specifically, BNs are probabilistic graphical models which embed data, expert opinion or a combination of the two into an intuitive graph thereby enabling reasoning given inherent uncertainty.[20] The basic structure of a BN (Fig. 1A) consists of nodes, each representing a variable, and arcs (arrows) which connect nodes in a causal relationship.[21] Underlying this intuitive and transparent structure are Bayes’ theorem and Bayesian statistics which allow for calculating conditional and joint probability distributions across many variables within the network.[22, 23] The BN allows for both data and domain knowledge to be packaged into a single model that can yield insights about real-world probabilities, can be updated with new data or opinion over time and can tolerate missing information.[13, 20, 21] Here, we describe a BN which both assesses risk of underlying primary immunodeficiency and provides clinical guidance. (Fig. 1B) Our network model was constructed by an expert immunologist but the parameters (i.e. conditional probabilities) were learned entirely from data. Our model may be embedded within an electronic health record (EHR) via app-program interface (API) for the purposes of analyzing large-scale data in real time; alternatively, it may be clinician facing for data entry at the point of care to assess a given patient’s risk and guide diagnosis

**Fig. 1.**
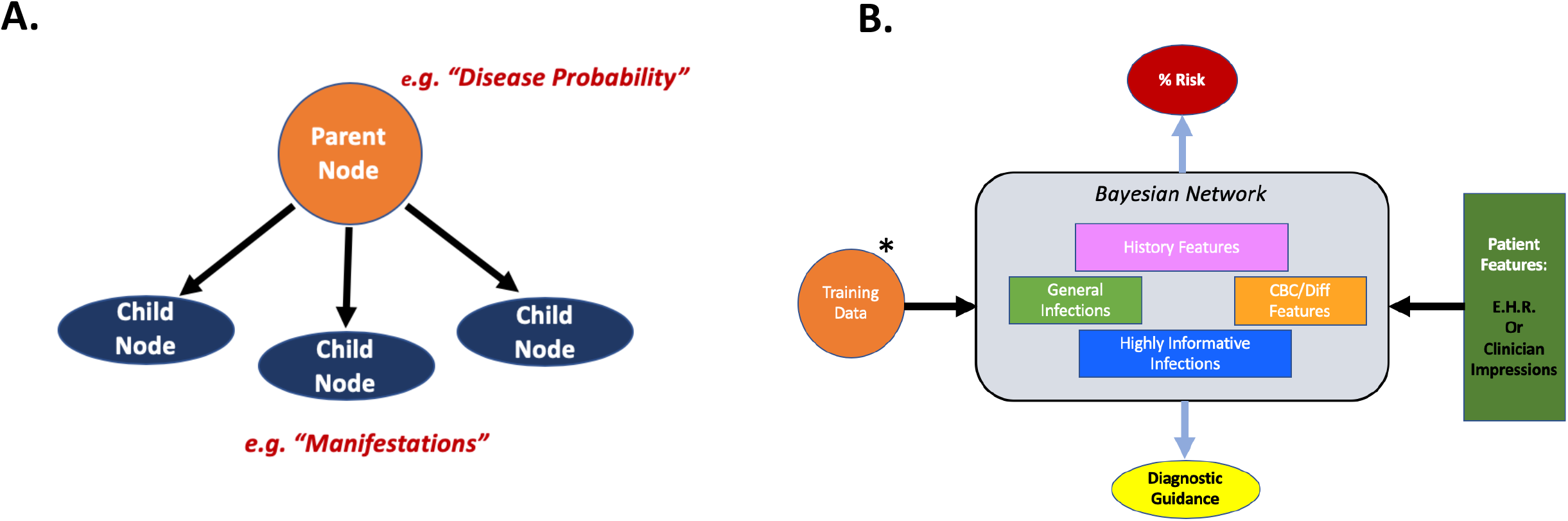
Bayesian Network Structure. **A**. An example Bayesian network with parent and child nodes connected by arcs. **B**. The general structure of our network components. Training data(*may be updated over time) and patient features allow for improving inference (i.e. probabilities) and structure over time making these networks dynamic. (EHR = Electronic Health Record)

## Results

### PI Cohort vs. Control Cohort

Demographic statistics for the PI and control populations are shown in Table 1. The proxy for social determinants of health (SDOH) and healthcare access via assessment of insurance coverage and type was similar between groups (PI Cohort-52% Private, 45% Public, 3% Unknown; Control Cohort-48% Private, 50% Public, 2% Unknown). Network feature nodes are shown in Table 2 and were all significantly different in the PI vs. control cohorts (p-value <0.0001).

**Table 1:** Demographic Features of the Cohort.

|  | PI Cohort<br>(n = 1762) | Control Cohort<br>(n = 1698) |
| --- | --- | --- |
| Mean Age (yrs) | 8 ± 4 | 7 ± 5 |
| Sex |  |  |
| Male | 1001 (56.8%) | 891 (52.5%) |
| Female | 761 (43.2%) | 807 (47.5%) |
| Ethnicity: |  |  |
| Caucasian | 793(45%) | 552(33%) |
| Hispanic | 606(35%) | 601(35%) |
| Black | 199(11%) | 281(17%) |
| Asian | 89(5%) | 91(5%) |
| Unknown | 75(4%) | 173(10%) |

**Table 2.**
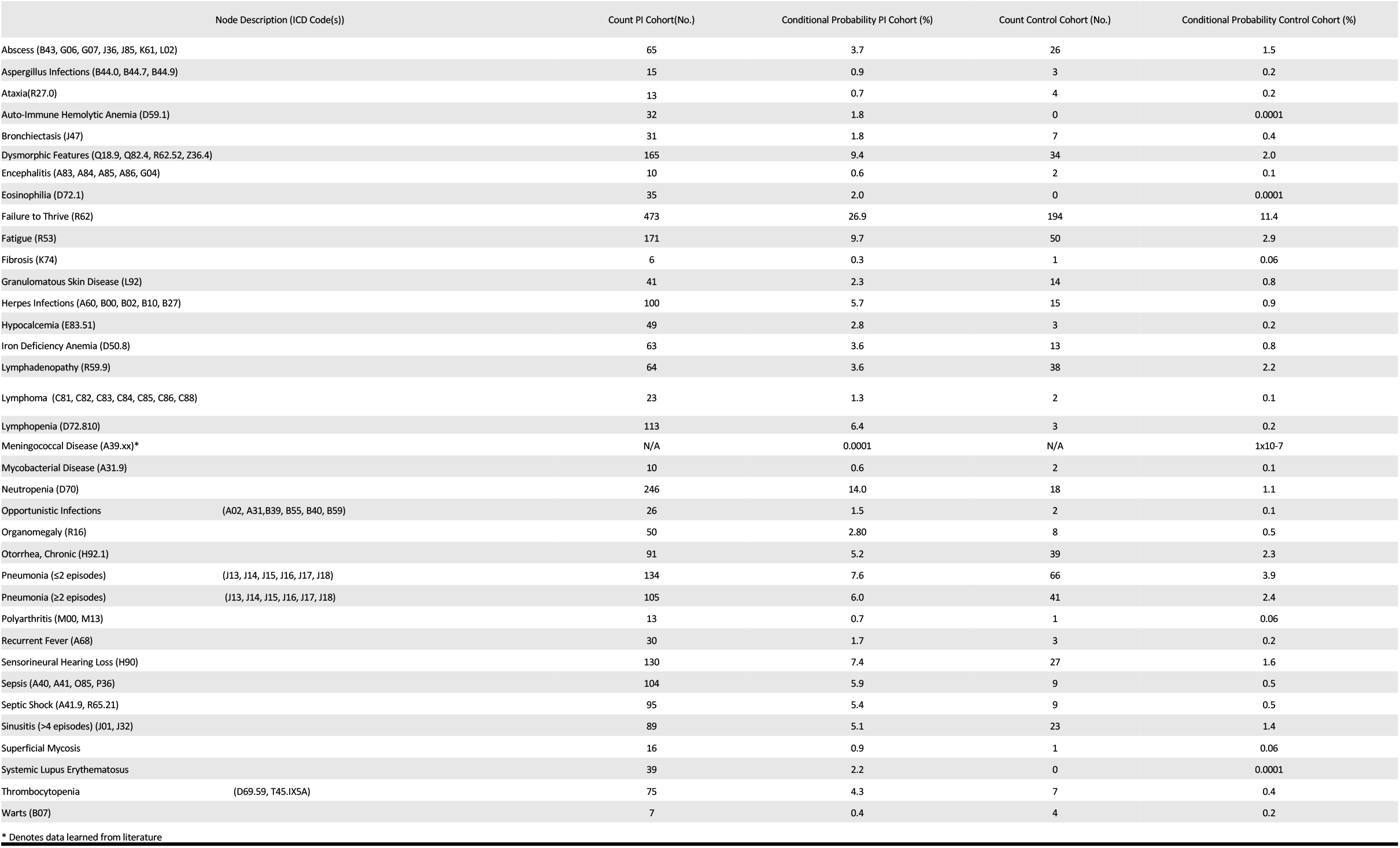
Network features with corresponding counts and conditional probabilities for each cohort.

### Risk Calculation (Cohort vs. Controls)

Performance of the BN on our validation cohort is shown in Figure 3. Individual risk calculation is displayed for the immunodeficient and control patients (Fig. 3A) as a single assessment for a given patient at the time of chart review. Classification performance is shown in Figure 3B and displayed as a receiver operator characteristic curve (ROC). The ROC analysis predicted best model performance at a risk cutoff of >5.5% (Sensitivity = 87%; CI(77%-93%) and Specificity = 91%; CI(82%-96%)). The corresponding BN performance measures were subsequently calculated as reported in Table 3 for risk calculation >5.5%.

**Table 3:** Network Performance (Risk Calculation)

|  | Score |
| --- | --- |
| Specificity | 91% |
| Sensitivity | 87% |
| PPV | 91% |
| NPV | 87% |
| Accuracy | 89% |
| F1 | 88% |

PPV-Positive Predictive Value; NPV-Negative Predictive Value; F1=F1 Score

**Fig. 2.**
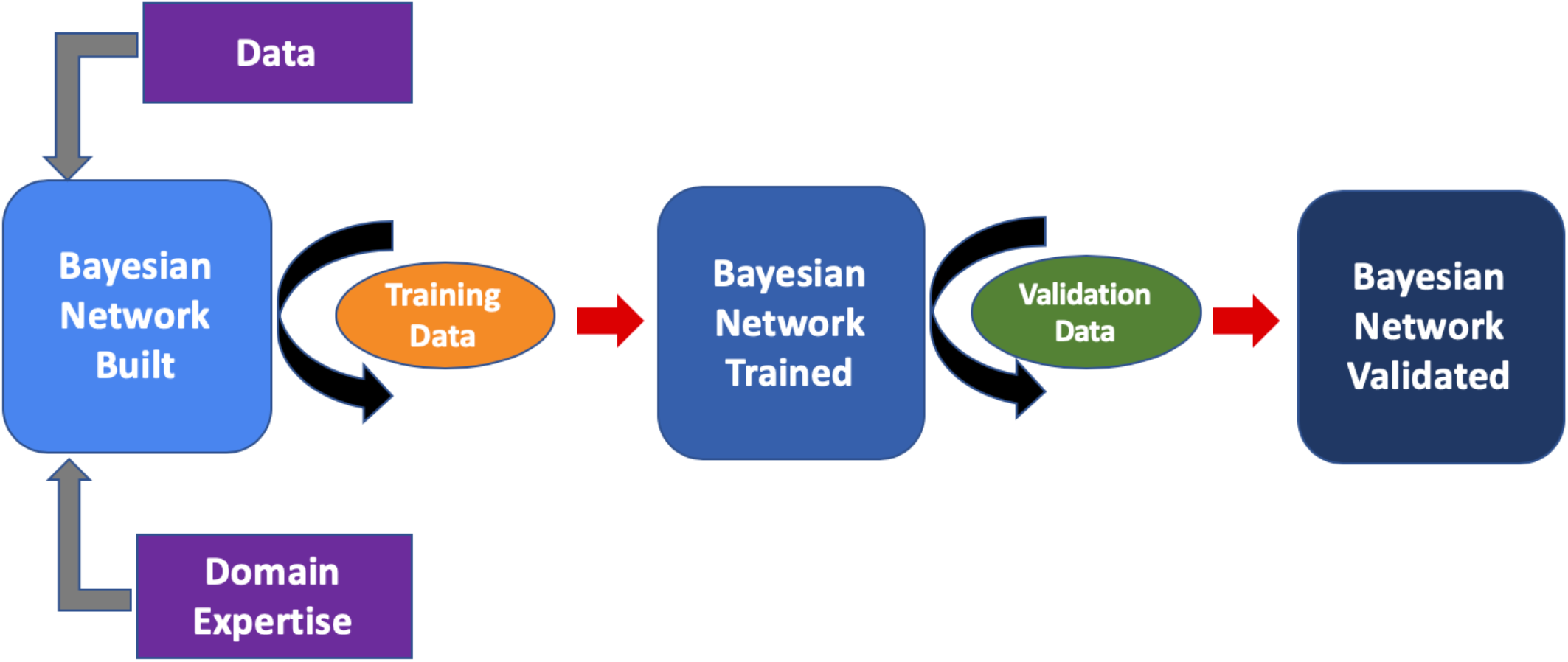
Analytics Plan. Network construction, training and validation scheme. Here, 100 patients (50 PI and 50 Control) were used as training data and 150 (75 each cohort) were used as validation data.

**Fig. 3.**
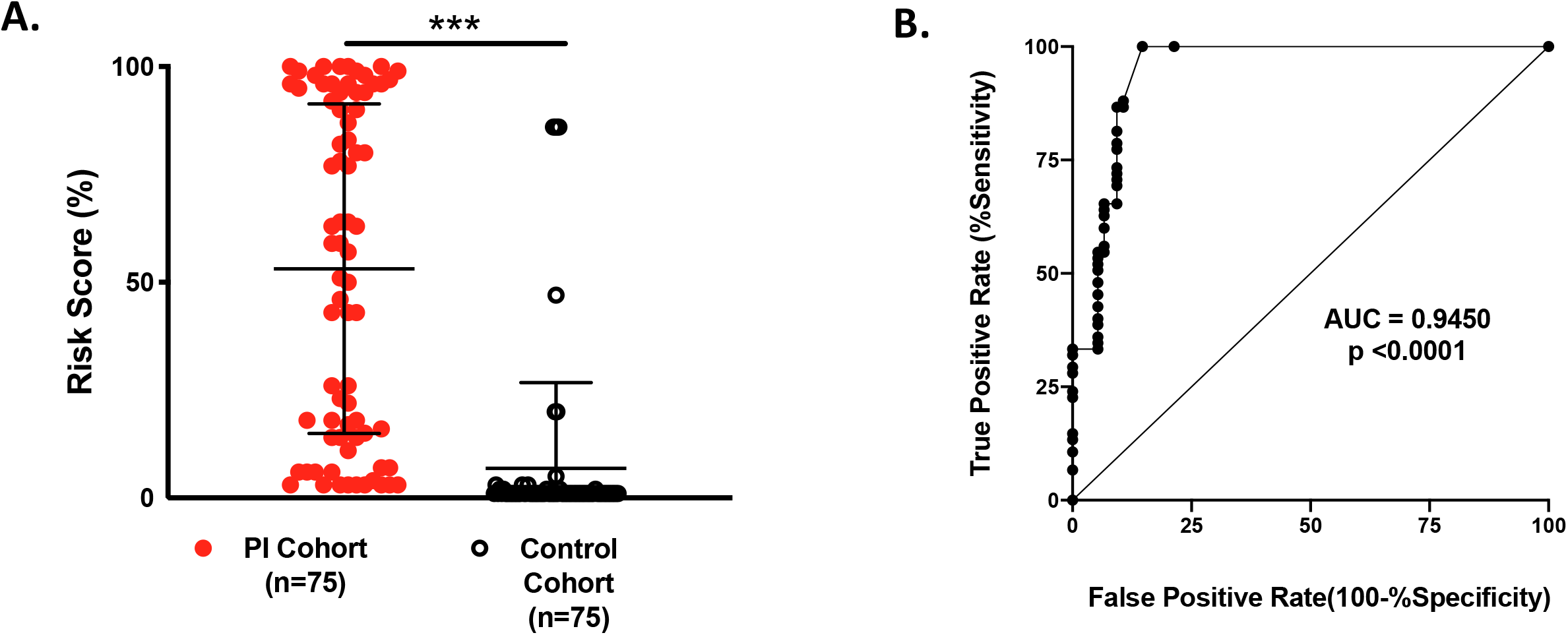
Network Validation. Validity testing of our BN for individual patients from the PI and Control cohorts. **A**. Mean risk scores between the two populations were significantly different (53% vs. 7%; p <0.000001). **B**. Network performance as calculated by AUROC (Area under Receiver Operator Characteristic Curve) where an AUROC of 1.0 represents the ability of a model to discriminate between classes 100% of the time.

### Prescriptive Output – Directing Clinical Management

Individuals within the PI validation cohort (n = 75) had 20 distinct PI disorders spanning the first 8 IUIS category tables. (Fig. 4A) Within the validation cohort, most had a Table 1/Table 2 (T/CID) related disorder (n = 32; 43%) or a Table 3 (PAD) related disorder (n = 26; 35%). Of the remaining disease categories, Table 4 (PIRD) comprised 1%(n = 1), Table 5 (PD) comprised 8%(n = 6), Table 6 (ID) comprised 3%(n = 2), Table 7 (AID) comprised 4%(n = 3) and Table 8 (CD) comprised 1%(n = 1).

**Fig. 4.**
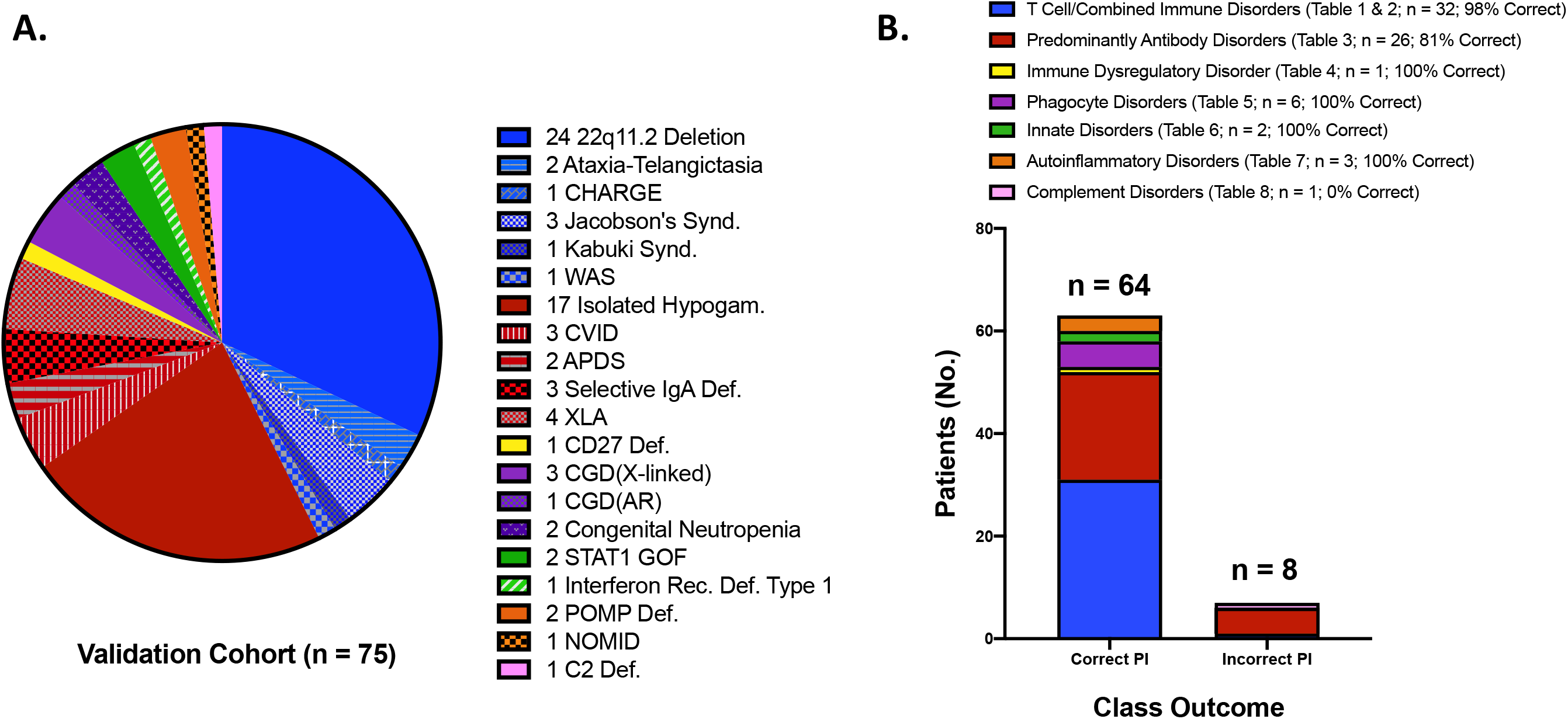
Cohort Features & Network Outcomes. **A**. Validation cohort disorder spectrum. The IUIS groupings are clustered according to color (i.e. Blue=T/CID; Red=PAD; Yellow=PIRD; Purple=PD; Green=ID; Orange=AID and Pink=CD. **B**. BN performance for classifying each IUIS category and overall outcome. The legend displays category number and accuracy for our BN prediction. NOTE-3 patients were not included here since insufficient input data were available and a class outcome could not be defined by the model. (Abbreviations: CHARGE-coloboma, heart disease, atresia of choanae, restricted growth, genital and ear abnormalities; WAS-Wiskott-Aldrich Syndrome; CVID-Common Variable Immunodeficiency; APDS-Activated Pi3K Delta Syndrome; XLA-X-linked Agammaglobulinemia; CGD-Chronic Granulomatous Disease; STAT1 GOF-Signal Transducer Activator of Transcription 1 Gain of Function; POMP-Proteasome Maturation Protein; NOMID-Neonatal Onset Multisystem Inflammatory Disease.)

Model performance for predicting each patient’s top 2 most likely IUIS disease categories is shown in Figure 4B. The BN was most successful in diagnosing patients with T/CID and phagocytic disorders. It was moderately effective in classifying antibody deficient patients appropriately. Additionally, the BN accurately classified patients with immune dysregulation, innate disease and autoinflammatory disease, but failed to classify the one complement disorder patient in our cohort. Overall BN model accuracy for determining patients across any studied IUIS category was 89%. It was 86% accurate if patients with table 1-3 disorders were omitted (n = 17).

### Bayesian Network Functionality & Usability

Data flow though the BN occurs once patient information is instantiated by an end-user (via our web portal for example - https://bcm-demo.bayesfusion.com/dashboard.html?id=3gml3vd510t3qhb366xg3uevp) or by analyzing diagnostic codes by EHR automatic feed via an API.(Supplemental Fig. 1B & C) The native structure is shown in Panel A; however, one can see the outputs for “Risk” and IUIS Category change depending up on data input. As displayed in Supplemental Figure 1B.1 & 1B.2 a patient’s data with X-linked agammaglobulinemia (XLA) and associated clinical features are instantiated. The BN then provides an updated risk prediction and a prescription about the most likely diagnostic category (i.e. PAD) which can then inform the initial immunological workup. Supplemental Figure 1C.1 & 1C.2 shows information flow for a patient with chronic granulomatous disease (CGD) whose clinical findings are instantiated. Here risk is again updated from baseline and the prescriptive component selected PD as the top predicted IUIS category. A schematic of the proposed workflow is shown in Figure 5.

**Fig. 5.**
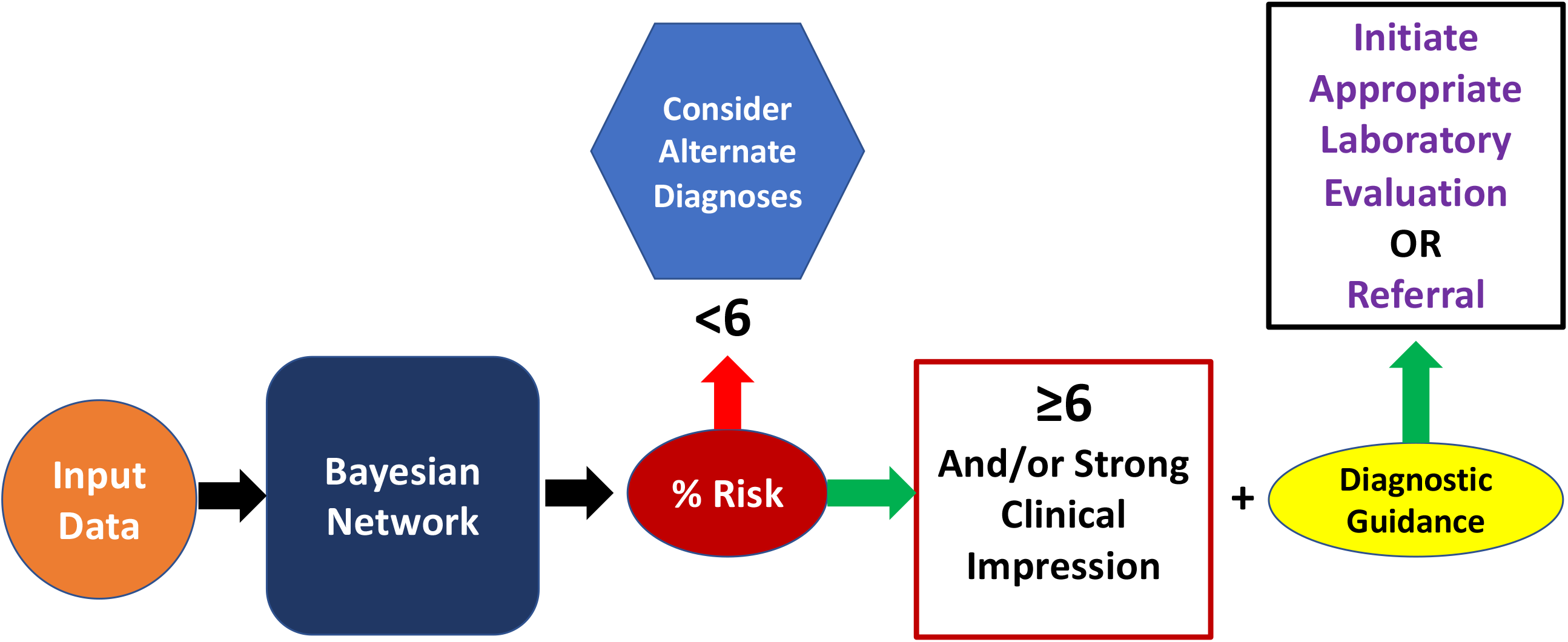
Workflow Model. The proposed workflow for our model. Here, an end-user or EHR data feed can provide inputs via clinical impressions or diagnostic codes. The BN calculates a risk score which can subsequently be acted upon. It is important to note that it is the risk score and clinical impression should be taken together, which guide subsequent evaluation and management.

## Discussion

Knowledge about the biology of PI is expanding rapidly; however, overall diagnostic rates have not appreciably improved.[2, 3, 5] Therefore, novel disease-detection methods are needed such as digitizing relevant phenotypes and leveraging health information technology functionality. Here we demonstrate that concepts related to PI can be learned from readily-available, EHR-mined, structured data and embedded within a transparent (Fig. 1B) machine learning model that accurately provides both risk assessment and clinical guidance(i.e. ICDCs; Table 2, Figure 2). The model, a BN, allows for concept linking driven by domain expertise thereby enabling information flow in a rational manner for disease detection.[6, 14, 24, 25] This model can be deployed in almost any clinical setting and may learn over time as population specific conditional probabilities are accumulated with ongoing accuracy refinement.[26, 27]

Our model performed well on an unseen validation cohort of 75 patients with PI possessing a diverse range of diseases across the IUIS spectrum. (Table 3, Figure 3 & Figure 4) The AUROC (0.945; p < 0.0001, Figure 3B) suggests robust model classification of PI patients vs. control individuals. Training of the model consisted of analyzing 50 PI patients and 50 controls to assess performance which allowed us to identify gaps in concept relationships (Figure 2). Figure 3A shows considerable scatter across risk scores for the PI validation cohort; however, the mean group score was significantly different from that of the controls (53.1±38.1 vs. 6.9±20; p < 0.000001) providing evidence that our model’s inference about PI is sound. Importantly, our age-matched and SDOH-similar control group consisted of patients with varying medical complexity themselves but not PI. Diagnoses among this group included cystic fibrosis, trisomy 21, acute lymphoblastic leukemia, solid organ cancers, asthma, complex congenital heart disease and complex genetic disease. Therefore, accurately distinguishing PI from our controls is likely a more challenging task than classification against the general pediatric population. We take this as evidence of real-world model fitness and utility for our BN.

In addition to risk prediction, our model provides a prescriptive outcome to facilitate appropriate diagnostic evaluations and initiate referral if needed (Figure 4 & Figure 5). The BN’s ability to direct a diagnostic approach is two-fold (Figure 5). First, a clinician’s threshold for testing should be triggered by their clinical impression and a risk score ≥6%. The next question, “what do I do now?”, can be answered via guidance about the most likely IUIS category. Given noted phenotypic heterogeneity among PI disorders, we assumed the initial predicted IUIS category would not be sufficiently inclusive; thus, we built our BN to predict the top 2 IUIS categories for each PI patient.[28] Using this strategy, our model was able to accurately define this class 89% of the time for the PI validation cohort (Figure 4). In 3 cases, the BN was unable to provide clinical guidance owing to lack of sufficient information. This scenario results in an equal probability prediction across IUIS categories and underscores the diagnostic uncertainty when scant patient information is available.

The prescriptive output of our model has utility for the primary care provider and clinical immunologist. Our expectation is that the provided decision guidance will inform clinical encounters by expediting appropriate testing and accurate diagnosis, assuming availability of recommended immune-diagnostic testing.[29] For the generalist or clinicians who are not generally focused on PI patients, we envision enabling early initial testing and referral to improve diagnostic rates. For the expert clinical immunologist, having relevant results in hand at the time of initial encounter should facilitate early implementation of best treatment practices and drive optimal outcomes for patients.

From an epidemiologic standpoint, our BN’s best predicted risk score cutoff of ≥6% is interesting in that it aligns well with our previous work in calculating a risk vital sign for PI.[4] There, analyzing a different population, we found 1% of the general population to be at medium-high risk for PI and subsequently ∼5% of this group had PI or a concerning infectious diagnosis in the following year.[4] These results suggest that the prior probability of disease (prevalence) approaches 5% for individuals deemed to be of medium-high risk. Thus, all healthcare providers may expect approximately 1% of their patients to be at risk for PI. These individuals must then be distinguished from individuals with actual disease further reinforcing the importance of considering this vulnerable patient population amongst anyone with infections.

Here, we hypothesize that informatic methods may inform the diagnostic process by combining disease-specific features and epidemiologic data to enable diagnosis. Such models require computational transparency, good performance and should extend optimal digital health workflows to align with clinical decision support (CDS) best practices.[30] Predictive analytics allows for discrimination about what might happen in a given clinical scenario; whereas, prescriptive analytics focuses on what one should do about the prediction.[31] This BN packages these attributes and presents a dual output for each patient to the user. Given reported concerns about time pressure for healthcare providers, delays in diagnosis, increased costs and poor outcomes we hoped to address these obstacles with our model en route to lowering the bar for PI diagnosis.[2, 7, 32] Lastly, we wish to underscore that our view of AI’s role in healthcare is that it should be rigorously validated but dutifully implemented to augment clinical decision making and extend clinical efforts for making use of big data in a way that serves the patient and provider.[33, 34] With this perspective AI/machine learning aids the clinician and makes health information technology more useful.

## Limitations & Future Directions

While our model performed very well with the validation cohort, we need to test it prospectively on larger numbers of patients. Also, it will be helpful to have a larger number of end-users provide feedback about our web-interface usability. Future work in this regard will focus on expanded testing and soliciting expert immunologist opinions broadly as they use the BN for patient assessments.

Another limitation of our study was the somewhat biased PI cohort with a large number of 22q11.2 deletion syndrome patients (n = 24). The model was very accurate in predicting their IUIS class, but some of these individuals may have been detected via TREC-based newborn screening. This bias is reflected in our overall PI population which contained many DiGeorge Syndrome patients cared for at our center. Additionally, we had limited representation of patients with disorders falling into the IUIS Tables 4, 6, 7 and 8. Patients with these disorders are less prevalent among PI as a whole; however, such information could be mined from registries or pooled from other centers and used to improve model fitness. Also, we decided to exclude inference about marrow failure syndrome and PI phenocopies (IUIS Tables 9 & 10) here; thus, patients with these disorders might not be detected with the current version of our network. We can track model performance prospectively about patients with such disorders and easily modify the network as needed.

Lastly, we plan to investigate additional variables that can be added to further improve network performance. Our BN performs very well in validation with only 36 nodes; however, it is likely that additional informative features will drive further improvement.

## Conclusions

Machine learning may facilitate diagnosis of patients with PI. Combining domain expertise and readily available EHR structured data with well-defined machine learning models provides an effective tool for risk assessment of PI.

## Methods

### Ethics Statement

This study was approved by the Baylor College of Medicine Institutional Review Board (H-38501). All patient data remained confidential and was retained in the Texas Children’s Hospital system.

### Cohort Analysis

Patients from Texas Children’s Hospital (TCH) with immunodeficiency were identified by having at least 2 ICD10 codes (ICDCs) entered at different time points (2008 – 2018) which was consistent with a primary immunodeficiency as categorized by the American Academy of Asthma, Allergy and Immunology (AAAAI; https://www.aaaai.org/Aaaai/media/MediaLibrary/PDF%20Documents/Practice%20Management/finances-coding/ICD-10-Codes-Immunodeficiencies.pdf). The PI cohort was comprised of 1762 individuals. (Table 1) In contrast, control data were specified by an age matched set of patients from TCH who did not harbor one of the PI codes and consisted of 1698 individuals. This was termed the “Control Cohort”. For each cohort, all ICDCs over 24 months were extracted from the TCH EHR (Epic Clarity Database). Individual, unique ICDCs were compiled for each cohort, counted, normalized and assessed for differences between groups. Relevant ICDCs were defined as having a significantly increased frequency among PI patients and associated relevance to PI based upon domain expertise. Features for construction of the BN were derived from relevant ICDCs with pertinence to human immunological disease as determined by an expert clinical immunologist (NLR). (Supplemental Figure 1)

### Prior Probability Calculations

We calculated prior probabilities for 36 different feature nodes for both PI the and Control populations. Each feature had corresponding ICDCs as listed in Table 2 and was representative of the node concept.[35] Prior probabilities were calculated by taking the total number of occurrences for each ICDC of interest and dividing by the total number of patients in that specific cohort (either PI or Control). These conditional probabilities (i.e. fraction of individuals with the condition given PI or given non-PI) were then used to create the Bayesian Network as described below. When there were no occurrences of a given clinical entity among controls, we arbitrarily set the conditional probability to be 0.0001% since a prior value of “0%” or “100%” does not accurately reflect real-world uncertainty. Learning prior probabilities for the “meningococcal disease” node general risk differed from the rest in that it was derived from literature rather than from data; additionally, data about patients receiving complement inhibitor therapy were used as a surrogate for meningococcal disease risk in PI.[36, 37]

### Bayesian Network Construction

Network structure was designed to facilitate clinical decision making derived from information obtained from a thorough history, physical examination and complete blood count (CBC/differential) without need for advanced immunological testing. The BN described was constructed using GeNIe Modeler available free of charge for academic research and teaching use from BayesFusion, LLC (http://www.baysefusion.com/). Feature nodes were created and added from learning as described above. In total 36 feature nodes were used (10 history/physical examination, 5 laboratory, 5 general infections, 16 highly informative infection/condition) which corresponded to 79 distinct ICDCs. (Figure 1B, Supplemental Fig 1, & Table 2) Contained within each node are the conditional probabilities of finding that variable in the PI cohort (“med-high risk”) and within a general pediatric population(“low risk”). We assumed the general population overall *risk* to be 1% for PI. This threshold was determined experimentally from our prior work where 2188 of 185,892 individuals (1.2%) were deemed at medium-high risk for PI.[4] Our network structure, dynamic information flow and intuitive dashboard can be found at https://bcm-demo.bayesfusion.com/dashboard.html?id=3gml3vd510t3qhb366xg3uevp

The BN structure was designed such that all 36 diagnostic variables/features are connected to the risk node. Risk is then calculated by employing Bayes’ Theorem for single or multiple conditions as features are instantiated for a given patient. Feature nodes were also connected via arrows (edges) to each of 8 IUIS disorder categories described as: T-cell/Combined (T/CID), Predominantly Antibody(PAD), Immune Dysregulatory(PIRD), Phagocyte(PD), Innate(ID), Autoinflammatory(AID) and Complement(CD). We combined Tables 1 & 2 from the Expert Committee report into one category and did not include bone marrow failure syndromes or PI phenocopies. This resulted in 7 IUIS disorder network nodes. Node connections were created based upon their contribution to a phenotype for that category as expected by domain expertise or as described in Tangye et al.[3] (Supplemental Fig. 1) Arrow directions indicate dependency (e.g. moving in the direction of the arrow one can say “*this* is dependent upon *that*”). For a given patient, IUIS category likelihood ranking was calculated by summing individual conditional probabilities among the present feature nodes. The output IUIS categories with the highest “yes” probability suggest the most appropriate starting point in the diagnostic workup for an individual patient. The probability output shown is compared to the general population; therefore, magnitude change from baseline is more important than the absolute percentage shown.

### Bayesian Network Training & Validation

From our immunodeficient and control cohorts, 250 patients (125 disease, 125 control) were randomly selected to train and validate the network. For the training and validation cohorts, each patient’s clinical and relevant laboratory history was confirmed by medical record chart review. Additionally, we assessed SDOH via analysis of insurance type as a proxy to normalize for health care access across the cohorts. Features were determined and entered into the BN for calculation of risk and IUIS category ranking. (Figure 2) For PI cohort training and validation, we excluded patients with a diagnosis of severe combined immunodeficiency (SCID) since they should have been detected asymptomatically via newborn screening. Similarly, patients with secondary immunodeficiency were excluded from the PI group but not from the control group.

After initial model build(i.e. node selection, probability embedding and arc-setting), we trained our BN with 50 PI patients and 50 controls*(“training data”*; Fig. 2). This allowed for tuning of the BN, adding additional relevant features and provided insights about node connections. Once the network was restructured, we performed validation testing with 75 previously unseen patients from each cohort*(“validation data”*; Fig. 2) Members of the validation cohorts had similar ages (mean control age = 8 ± 5yrs; mean PI age = 8 ± 3 years). Network performance was assessed by receiver operator characteristic (ROC) curve analysis on the validation cohort.

### Statistical Methods

Descriptive statistics for the PI and control cohorts were determined using Microsoft Excel and are shown in Table 1. Significance testing and the ROC curve analyses were conducted in Prism GraphPad version 8 (https://www.graphpad.com).

## Data Availability

Patient protected health information (PHI) is not available; however, all clinical data used to generate inference by the Bayesian network is available. Most relevant data from our study are included in the supplementary materials. Additional data availability may be made available from the corresponding author on reasonable request.

## Acknowledgements

We wish to thank the Jeffrey Modell Foundation for grant funding (JMF Translational Grant-58293-I). We are also grateful to our colleagues who contributed clinical care to patients whose data was analyzed here. We acknowledge Tomasz Sowinski of BayesFusion (https://www.bayesfusion.com/) for access to the Bayesian network modeling tool made freely available to academicians and Mr. Sowinski’s substantial work with creating the web-based dashboards.

## Author Contributions

NLR conceived the study, wrote the manuscript, had access to all data, oversaw data analysis, prepared the manuscript and helped design the web dashboard. GC performed statistical analyses and contributed to manuscript construction and review. TM, LW, AK, LMN, FOS and IKC extracted and contributed data and provided critical review of the manuscript. KR provided critical review of the network methods, contributed to the study concept design and manuscript review.

---

**Supplemental Table 1:** Network Nodes and Associated ICD10 Code Information.

| Network Node Label | ICD10 Codes & Descriptions |
| --- | --- |
| Abscess | B43 - Chromomycosis and Pheomycotic abscess; G06 - Intracranial and Intraspinal abscess and granuloma; G07 - Intracranial and Intraspinal abscess and granuloma in diseases classified elsewhere; J36 - Peritonsillar abscess; J85 - Abscess of lung and mediastinum; K61 - Abscess of anal and rectal regions; L02 - Cutaneous abscess, furuncle, and carbuncle |
| Aspergillosis | B44.0 - invasive pulmonary aspergillosis; B44.7 - disseminated aspergillosis; B44.9 - aspergillosis, unspc |
| Ataxia | R27.0 - Ataxia, unspecified |
| Auto-Immune Hemolytic Anemia | D59.1 - Other autoimmune hemolytic anemias |
| Bronchiectasis | J47 - Bronchiectasis |
| Dysmorphic Features | Q02 - Microcephaly; Q18.9 - Congenital Malformation of Face/Neck, Skeletal System; R62.52 - Short Stature; Z36.4 - Intrauterine Growth Restriction; Q82.4 - Ectodermal Dysplasia |
| Encephalitis | A83 - Mosquito-borne viral encephalitis; A84 - Tick-borne viral encephalitis; A85 - Other viral encephalitis; A86 - Not Elsewhere Classified; G04 - Unspecified Viral Encephalitis, Encephalitis, myelitis and encephalomyelitis |
| Eosinophilia | D72.1 - Eosinophilia |
| Failure to Thrive | R62 - Lack of expected normal physiological development in childhood and adults |
| Fatigue | R53 - Malaise and fatigue |
| Fibrosis | K74 - Fibrosis and cirrhosis of liver |
| Granulomatous disease of skin | L92 - Granulomatous disorders of skin and subcutaneous tissue |
| Herpes Infections | A60 - Anogenital herpes viral [herpes simplex] infections; B00 - Herpes viral [herpes simplex] infections; B02 - Zoster [herpes zoster]; B10 - Other human herpesviruses; B27.0 - Epstein Barr Virus/Mono |
| Hypocalcemia | E83.51 - Hypocalcemia |
| Iron Deficiency Anemia | D50.8 - Other iron deficiency anemias |
| Lymphadenopathy/ Generalized Enlarged Lymph Nodes | R59.9 - Enlarged lymph nodes, Unspec. |
| Lymphoma | C81 - Hodgkin lymphoma; C82 - Follicular lymphoma; C83 - Non-follicular lymphoma; C84 - Mature T/NK-cell lymphomas; C85 - Other specified and unspecified types of non-Hodgkin lymphoma; C86- Other specified types of T/NK-cell lymphoma; C88 - Malignant immunoproliferative diseases and certain other B-cell lymphomas |
| Lymphopenia | D72.810 - Lymphocytopenia |
| Meningococcal Disease | (A39.xx)*- Data learned from literature. For reference, relevant ICD10 codes include A39.xx. |
| Mycobacterial Disease | A31.9 - Mycobacterial infection, unspecified |
| Neutropenia | D70 - Neutropenia |
| Opportunistic Infections | A02 - Other salmonella infections; A31 - Infection due to other mycobacteria; B39 - Histoplasmosis; B40 - Blastomycosis; B55 - Leishmaniasis; B59 - Pneumocystis |
| Organomegaly | R16 - Hepatomegaly and splenomegaly, NEC |
| Otorrhea | H92.1 - Otorrhea |
| Pneumonia | J13 - Pneumonia due to Streptococcus pneumoniae; J14 - Pneumonia due to Hemophilus influenzae; J15 - Bacterial pneumonia (not elsewhere classified); J16 - Pneumonia due to other infectious organisms (not elsewhere classified); J17 - Pneumonia in diseases classified elsewhere; J18 - Pneumonia (unspecified organism) |
| Polyarthritis | M13 - Other arthritis; M00 - Pyogenic arthritis |
| Recurrent Fever | A68 - Relapsing fevers |
| Sensorineural hearing loss | H90 - Conductive and sensorineural hearing loss |
| Sepsis | A40 - Streptococcal Sepsis; A41 - Other Sepsis; O85 - Puerperal Sepsis; P36 - Bacterial Sepsis |
| Septic Shock | A41.9 - Sepsis (unspecified organism); R65.21 - Severe sepsis with septic shock |
| Sinusitis | J01 - Acute Sinusitis; J32 - Chronic Sinusitis |
| Superficial Mycosis | B36.9 - Superficial Mycosis, Unspec. |
| Systemic Lupus Erythematosus | L93 - Lupus Erythematosus; M32.9 - Systemic Lupus Erythematosus, unspecified |
| Thrombocytopenia | D59.59 - Other secondary thrombocytopenia; T45.IX5A - Adverse effect of antineoplastic and immunosuppressive drugs (initial encounter) |
| Warts | B07 - Viral Warts, etc. |

**Supplemental Table 2:** Immunodeficiency ICD10 Diagnoses for the PI Cohort (n = 1762)

| Diagnosis | Count (%) |
| --- | --- |
| Functional Disorders of Polymorphonuclear Neutrophils (D71) | 34 (1.9%) |
| Immunodeficiency with Predominantly Antibody Defects (D80) | 727 (41.3%) |
| Combined Immunodeficiencies (D81) | 27 (1.5%) |
| Immunodeficiency Associated with Other Major Defects (D82) | 314 (17.8%) |
| Common Variable Immunodeficiency (D83) | 76 (4.3%) |
| Other Immunodeficiencies (D84) | 872 (49.5%) |
| Other Disorders Involving the Immune Mechanism, Not Elsewhere Classified (D89) | 279 (15.8%) |

Note: Some patients had ICD10 codes associated with more than one of the diagnosis categories.

**Supplemental Fig.**
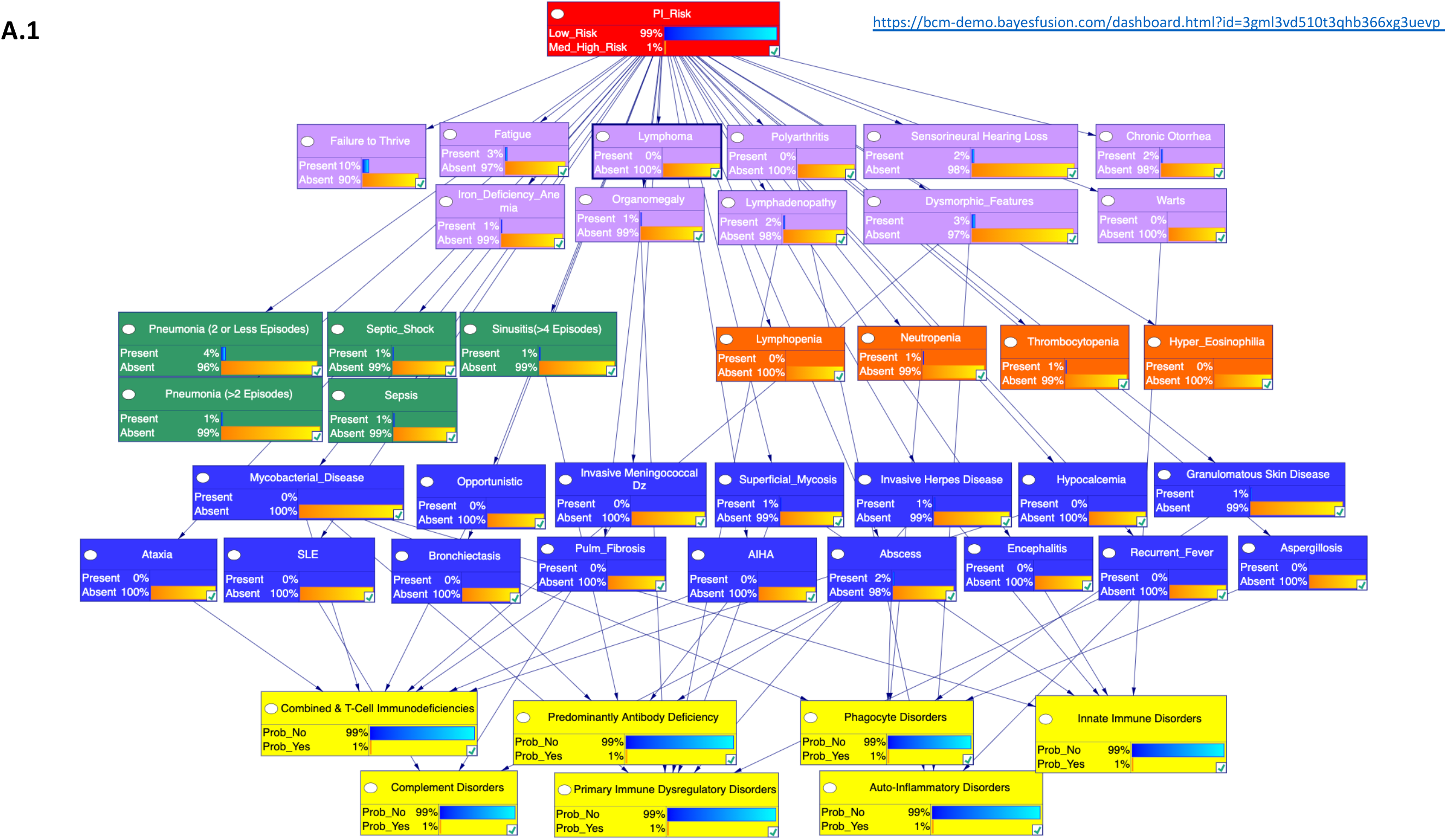
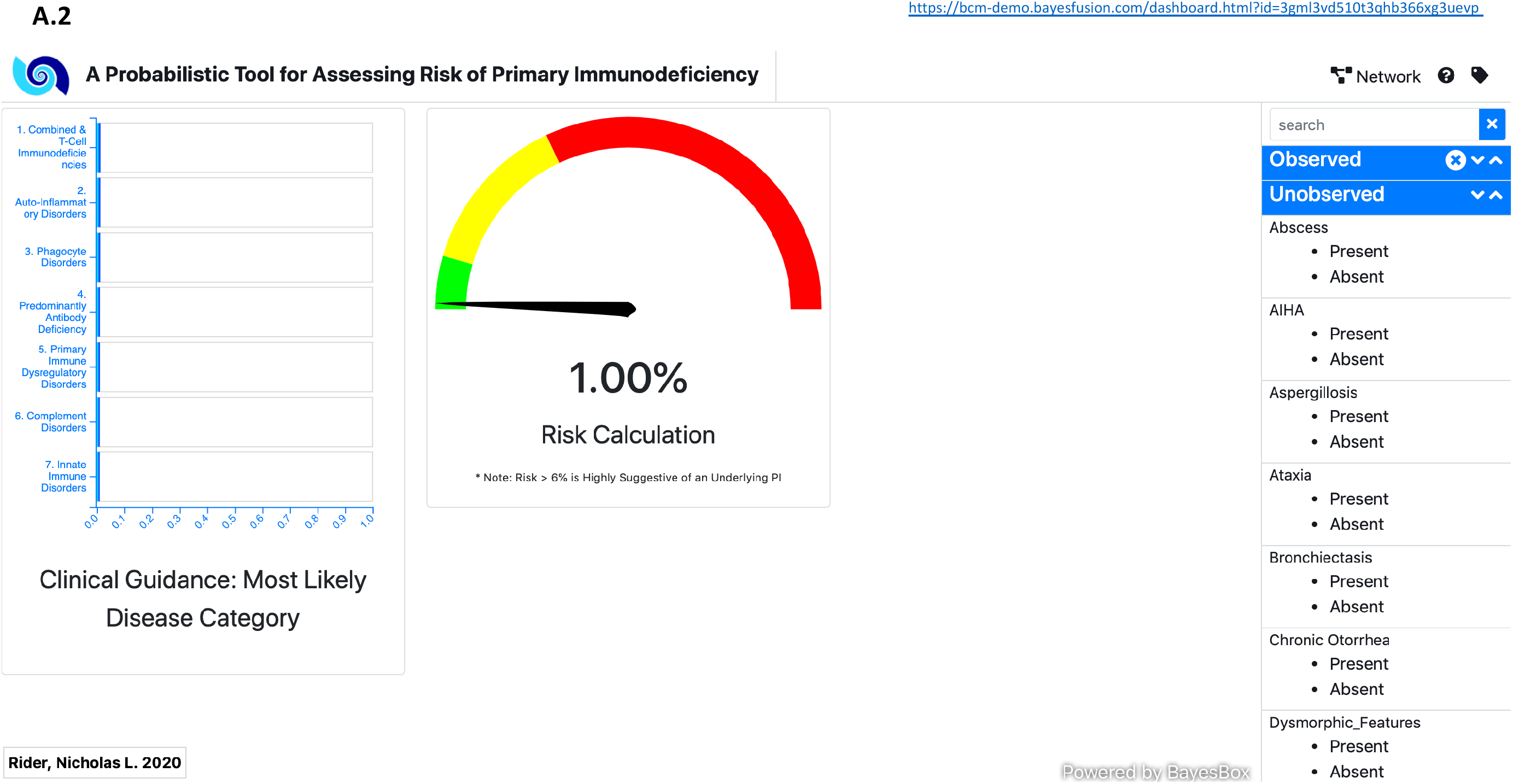
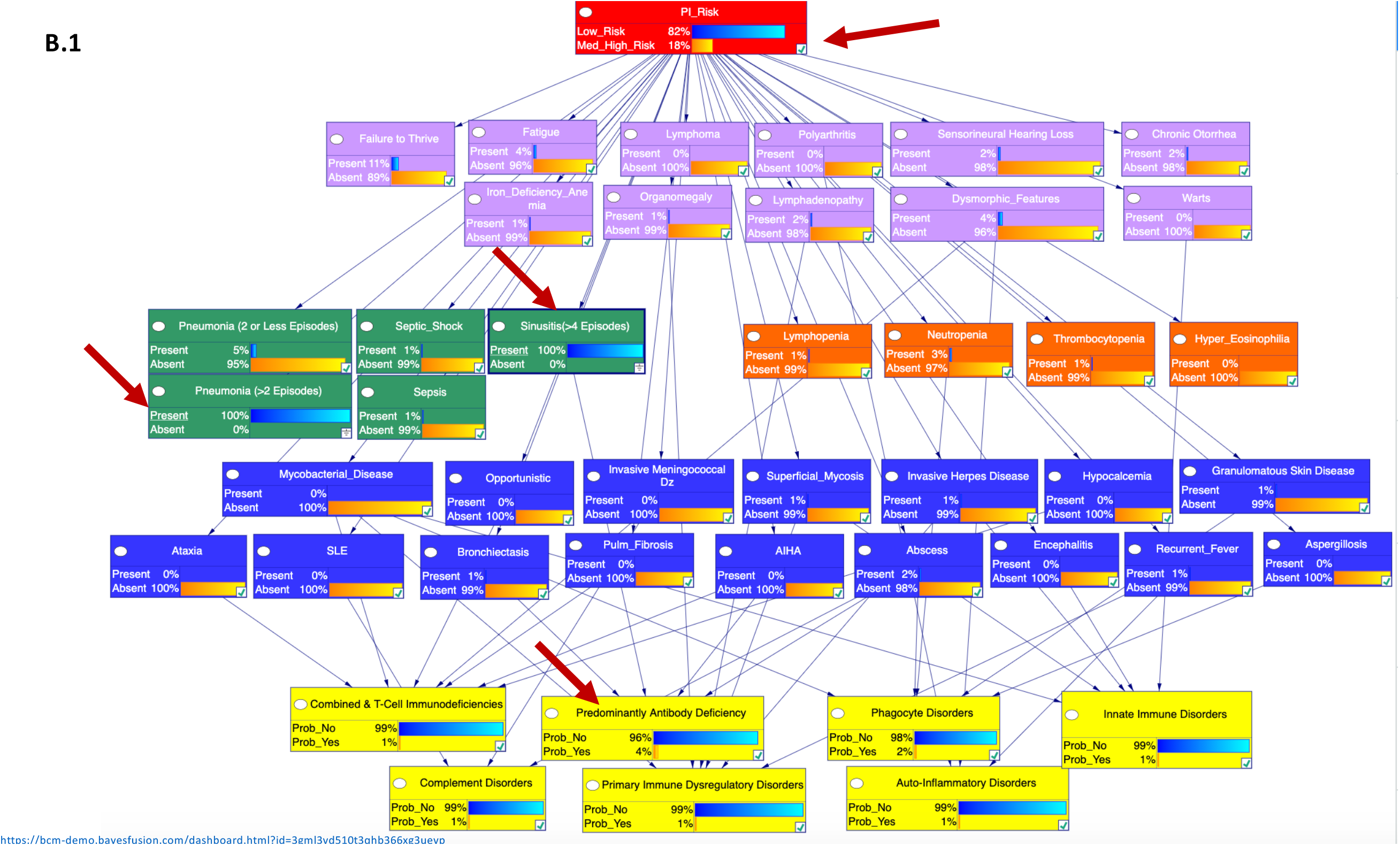
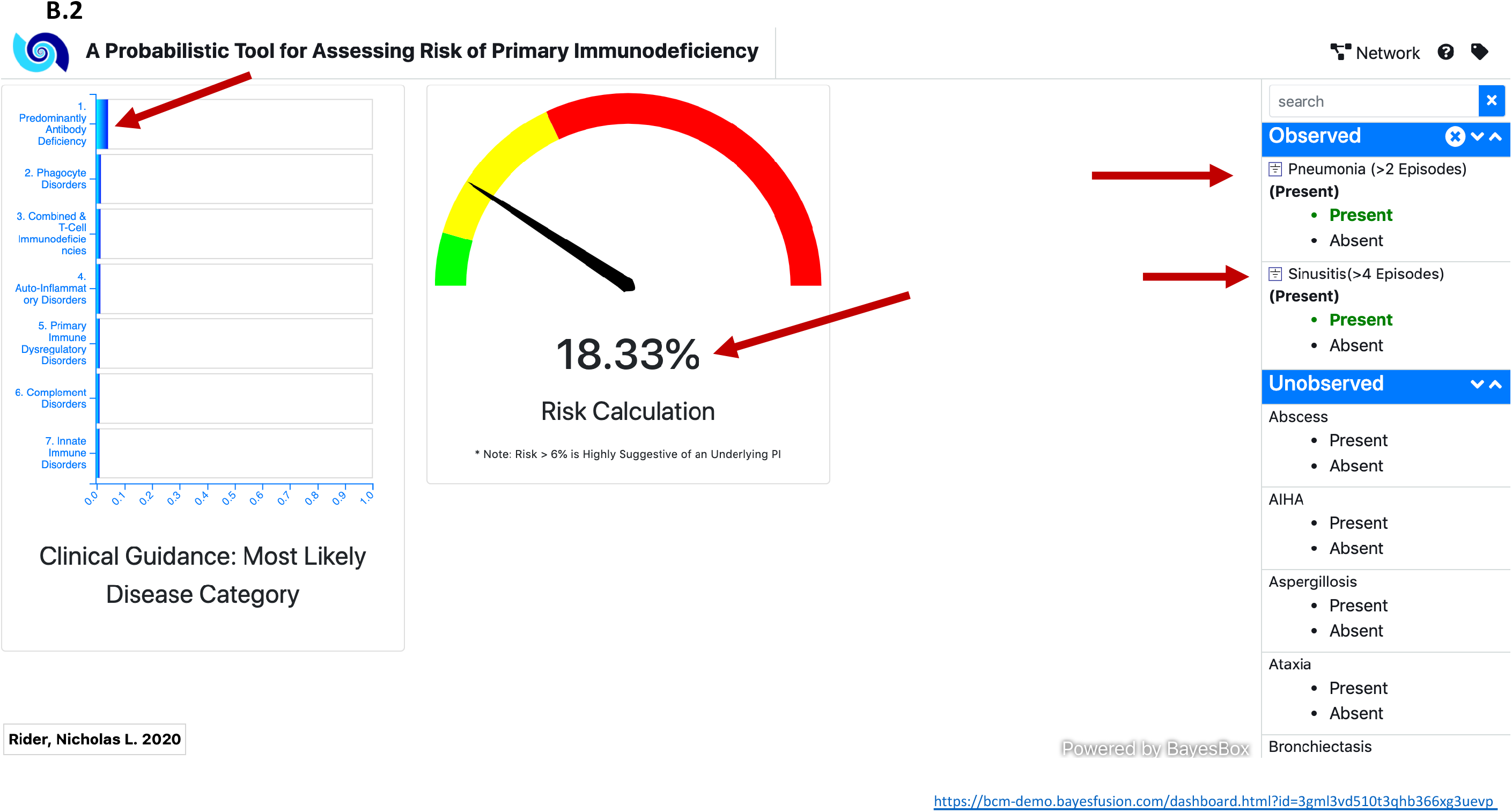
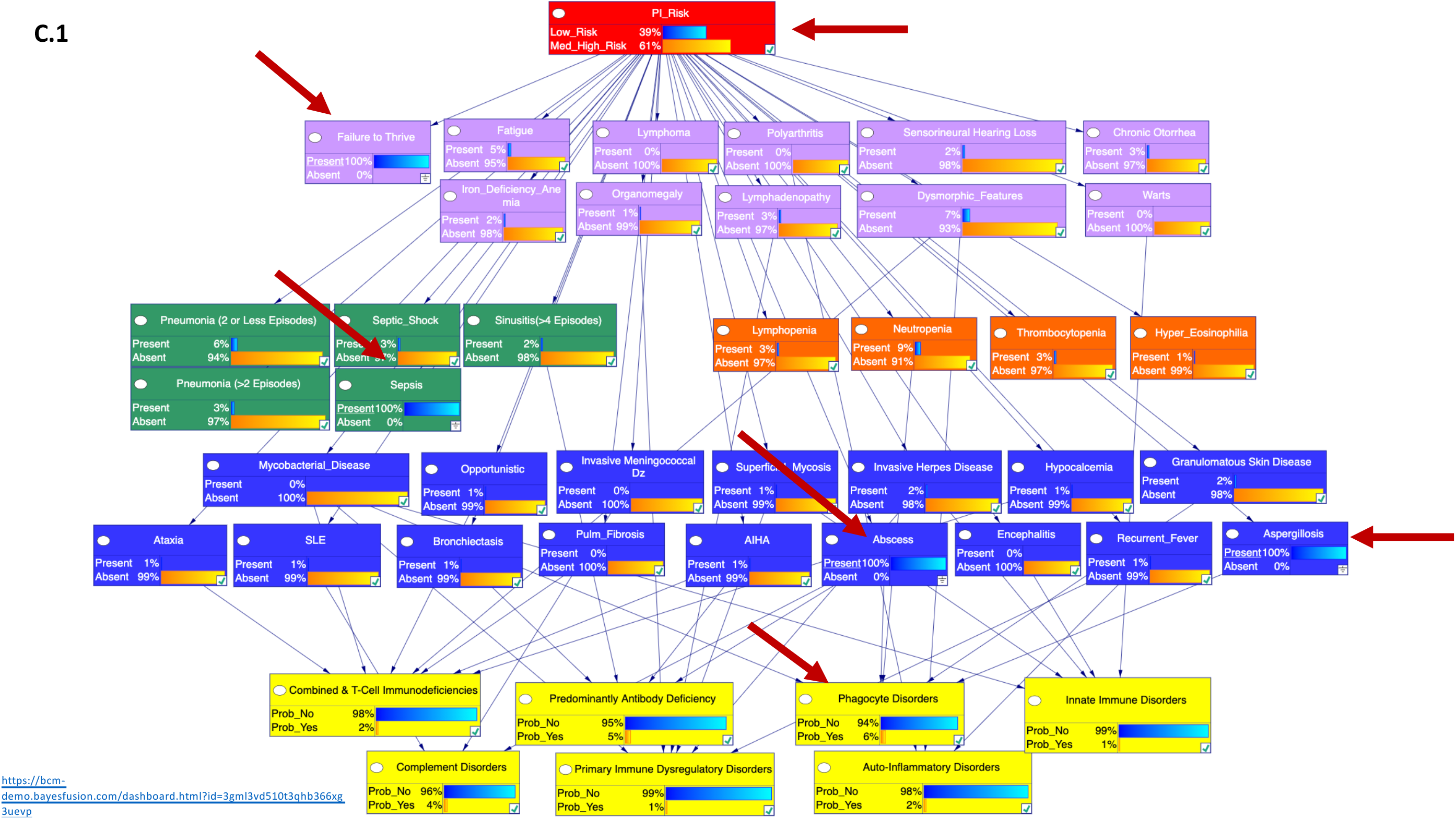
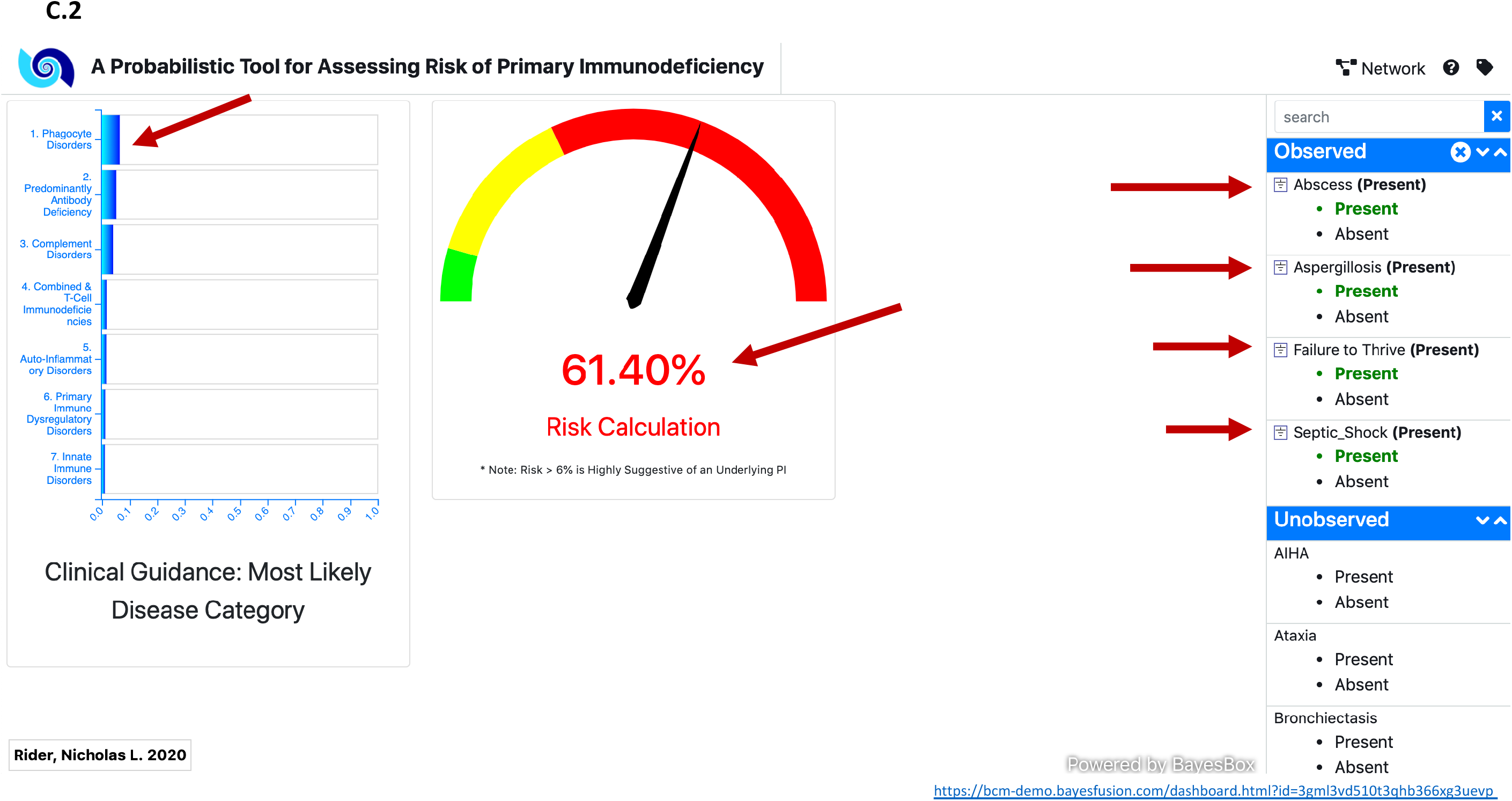
Web Interface-Network & Dashboard 1. **A**.**1** The native BN structure and low-risk probabilities specified are shown. Users can see background probabilities for low/high risk and select nodes here if desired. **A**.**2** Native dashboard interface. Users can select features along the right sidebar and see risk and top diagnosis prediction displayed on the left panels. **B**.**1** A case example of network information flow upon entering clinical data for a patient with X-linked agammaglobulinemia (XLA). **B**.**2** Dashboard output showing the most likely disease category (antibody deficiency) promoted to the top and risk calculation for the XLA patient. **C**.**1** A case example of network information flow for a patient with chronic granulomatous disease (CGD) and associated findings. **C**.**2** Dashboard output for the CGD patient displaying the most likely disease category (phagocyte disorder) promoted to the top with associated risk calculation. Note the arrows designating changes from the baseline probabilities as patient characteristics are entered.

